# Understanding the CLN2 Disease Journey: A Survey of Caregiver Experiences, Treatment Burden, and Treatment Perspectives

**DOI:** 10.64898/2026.05.31.26354557

**Authors:** Ineka T Whiteman, Katherine L Villa, Courtney M Spector, Jang-Ho J Cha, Amy Fenton Parker, Rebecca Ahrens-Nicklas, Angela Schulz, George J Yohrling

## Abstract

**Background:** CLN2 disease, Neuronal Ceroid Lipofuscinosis (NCL) type 2, is a rare, genetic neurodegenerative condition predominantly affecting children and characterized by seizures, progressive language and motor decline, vision loss, and premature death. Currently, the only approved disease-modifying therapy is the enzyme replacement therapy (ERT) Cerliponase alfa, administered fortnightly via intracerebroventricular infusion as a lifelong treatment. While ERT has been shown to slow motor and language decline, it is not curative and does not fully address all aspects of disease progression, including retinal degeneration. To better understand the lived experience of affected families, and perspectives on current and emerging treatments, we conducted a community survey of parents and caregivers of individuals with CLN2 disease.

**Methods:** A 25-question anonymous, voluntary survey was distributed through the US-based Batten Disease Support, Research and Advocacy (BDSRA) Foundation, and its international partner patient advocacy organisations via email, e-newsletters and social media. Eligible participants included current and bereaved parents or primary caregivers of individuals with CLN2 disease, regardless of treatment history. The survey explored treatment experiences and burden, unmet needs, and knowledge of and attitudes toward emerging therapeutic approaches, particularly gene-targeted therapies. Thematic analysis was performed on free-text responses.

**Results:** Ninety-eight respondents from 19 countries completed the survey, with the majority of participants from the United States and other predominantly English-speaking countries. Fifty-seven respondents reported current or prior use of ERT, with 94.7% (n=54/57) actively receiving treatment at the time of survey. ERT was perceived to provide greatest benefit for motor function and seizure control; however, respondents reported substantial treatment burden (mean burden score 4.8/7, n=66). Despite treatment availability, 94.9% of respondents (n=75/79) indicated a need for alternative therapeutic options and 94.8% (73/77) expressed interest in learning more about gene therapy. Overall, 72.4% (n=55/76) reported they were likely or very likely to consider participation in an investigational gene therapy trial. Key considerations included potential safety risks (57.9%, n=44/76), preclinical safety and efficacy evidence (54.0%, n=41/76), and whether ERT discontinuation would be required to participate (54.0%, n=41/76). Thematic analysis of 293 free-text responses identified substantial emotional, psychosocial and family impact, alongside treatment burden and persistent unmet need, hope for transformative therapies, and faith and resilience as prominent themes.

**Conclusion:** Despite the clinical benefits of ERT, CLN2 disease continues to impose substantial disease, treatment, and caregiver burden. Families expressed strong interest in emerging gene-targeted therapies while identifying safety, efficacy, treatment burden, and preservation of function as important considerations. Together, these findings demonstrate an ongoing need for therapies that can further modify disease progression, more comprehensively address both central nervous system and ocular manifestations of CLN2 disease and reduce treatment burden for affected individuals and their families.

## INTRODUCTION

CLN2 disease (OMIM #204500) is a rare, autosomal recessively inherited, neurodegenerative disorder belonging to a group of conditions called the Neuronal Ceroid Lipofuscinoses (NCLs), collectively called Batten disease. CLN2 disease is caused by pathogenic variations in the CLN2 gene that encodes the tripeptidyl peptidase 1 (TPP1) enzyme [1]. TPP1 deficiency leads to the abnormal lysosomal accumulation of ceroid lipofuscin, an autofluorescent amalgamation of lipoproteins, which contributes to cellular damage and death, particularly in the neurons of the brain and retina [2]. The “classical” late infantile onset CLN2 disease typically manifests between 2 and 4 years of age, often following an otherwise typical early childhood development. First presenting symptoms include language delay followed by, or in combination with, refractory seizures, ataxia [3–7], cognitive regression, progressive loss of speech and motor function, vision loss, painful movement disorders, and ultimately death, typically between 10 and 12 years of age when untreated [2,5,8,9]. Less commonly, individuals present with non-classical or “atypical” CLN2 phenotypes, characterised by later onset, variable symptomatology, and a more protracted disease course, including juvenile and adult-onset presentations [10–12]. These phenotypes are generally associated with residual TPP1 enzyme actvitity and have been reported at higher frequencies in some populations, including those in several Latin American and European populations [13].

Currently, the only regulatory approved disease-modifying therapy for CLN2 disease – and the only approved therapy available for any form of Batten disease - is the recombinant human TPP1 enzyme replacement therapy (ERT) Cerliponase alfa (Brineura®, BioMarin Pharmaceutical). In 2017, Cerliponase alfa was approved by the US Food and Drug Administration and the European Medicines Agency for the treatment of CLN2 disease and now has market authorisation in many countries around the world [14,15]. Cerliponase alfa is administered every two weeks directly into the cerebrospinal fluid (CSF) via intracerebroventricular (ICV) infusion using a surgically implanted ventricular catheter and subcutaneous (scalp) reservoir (typically a Rickham or Ommaya device) [16–18].

Cerliponase alfa demonstrates a significant, long-term slowing of motor and language decline in symptomatic classical and atypical CLN2 disease [16,19–22], with earlier initiation of treatment associated with more favourable outcomes [19–24]. Presymptomatic treatment can delay symptom onset [23–25], and ERT has also been associated with improved seizure control [22,26,27] and health-related quality of life [28]. Beyond ERT, current clinical management approaches for CLN2 disease are limited to symptomatic treatment and principles of palliative care[29,30].

Importantly, although ERT is shown to slow disease progression or delay onset in cases of presymptomatic treatment, it is not curative and does not address all aspects of disease progression. In particular, retinal degeneration and progressive vision loss continue despite ICV-delivered ERT [31–34]. Vision loss can severely impact quality of life for both affected individuals and their family, and emerging therapeutic approaches for CLN2 disease are currently targeting the stabilization or prevention of vision loss, including intravitreal ERT administration [35,36] and gene-based approaches [33,34,37,38].

Another notable limitation of cerebroventricular ERT treatment is the considerable burden associated with treatment administration, frequency and duration. Treatment is administered fortnightly on a lifelong basis, with each infusion requiring reservoir access via needle puncture in the scalp, and total infusion taking approximately 4.5 hours in addition to pre- and post-procedure requirements [16,39,40]. Reservoir material reportedly deteriorates after roughly four years requiring periodic surgical replacement of the device [18]. Additional complications reported in association with ventricular access devices include cerebrospinal fluid infection, perioperative or access-site swelling that precludes infusion, leakage or wound breakdown, and catheter blockage [17,22,40,41].

Beyond the profound clinical trajectory of the disease, there are substantial psychosocial, physical, and financial burdens, uncertainty surrounding treatment options, and intensive demands of ongoing care [42–45]. Despite these challenges, contemporary empirical data describing caregiver experiences in the era of ERT remain limited, particularly regarding the practical and psychosocial impacts of treatment and perspectives on an evolving therapeutic landscape. A deeper understanding of these lived experiences is essential for informing clinical care, optimising support services, and guiding future therapeutic development.

With nearly a decade of clinical experience with cerliponase alfa treatment since its first regulatory approval in 2017 [14][19] and several emerging therapies for CLN2 disease presently in development, the Batten Disease Support, Research and Advocacy (BDSRA) Foundation sought to better understand the lived experiences, concerns, and challenges of both current and bereaved caregivers within the CLN2 disease community. This study therefore aimed to characterise caregiver experiences of CLN2 disease and current treatment, and to explore knowledge of and attitudes toward emerging therapeutic approaches, particularly gene-targeted therapies.

## METHODS

The BDSRA Foundation developed a voluntary, anonymous online survey for the CLN2 Batten disease community using the SurveyMonkey software platform. The survey opened from 1 March to 29 March 2024. Invitations to participate were distributed by the BDSRA Foundation, and international partner patient advocacy organisations including BDSRA Australia, BDSRA Canada and Batten Disease Family Association (BDFA) United Kingdom. Recruitment occurred through public social media accounts and closed CLN2 family groups, direct email to the Batten disease community [46] and the BDSRA Foundation newsletter, Illuminator, which has more than 20,000 subscribers from at least 15 countries [47].

The survey was intended for all current and bereaved parents of children and young people affected by CLN2 disease. Where more than one individual with CLN2 disease was affected within a family, respondents were asked to complete a separate survey for each affected individual.

The survey comprised 25 questions addressing respondent and affected-individual characteristics, including country of residence, number of individuals with CLN2 disease in the family, current age, age at diagnosis, and the respondent’s relationship to the affected individual (see Supplementary Materials). Questions addressed the reported availability of ERT in the country of residence; treatment history, including age at initiation and reasons for discontinuation or non-initiation; symptom domains perceived to improve with ERT; and suggestions for improving the treatment experience. Additional questions explored caregiver and treatment burden, impacts on personal and family well-being, perceived need for alternative therapies, understanding of gene therapy, and willingness to consider participation in future regulated clinical trials.

The survey used conditional branching based on the child’s ERT history and current treatment status. Respondents who reported in Question 7 that their child had never received ERT were instructed to bypass Questions 8–12 and proceeded to Question 13, which assessed reasons for nonreceipt. Among respondents whose child had received ERT, those reporting current treatment in Question 9 bypassed Question 10, which assessed reasons for discontinuation, and proceeded to Question 11. ERT-experienced caregivers were asked about treatment initiation, current status, perceived benefits, opportunities for improvement, and treatment burden. All respondents subsequently re-entered a common survey pathway addressing the need for alternative treatments, broader caregiver burden, gene therapy awareness, and factors influencing potential clinical trial participation. Consequently, the number of respondents eligible for individual questions varied according to ERT history and current treatment status.

### Qualitative analysis

Free-text responses from 10 survey questions, comprising responses to open-ended questions and free-text fields accompanying structured questions, were analysed using thematic analysis informed by Braun and Clarke [53]. Responses were analysed as a single qualitative dataset while retaining the originating survey question as contextual information. An initial coding framework was developed primarily inductively from the data, while remaining informed by the clinical and caregiver-burden domains addressed by the survey. The preliminary framework comprised nine overarching themes with defined coding criteria.

Three authors independently piloted the coding framework using the same initial 20 responses. Each response was assigned one primary theme and, where applicable, up to two additional themes. The coders then compared their coding and discussed areas of disagreement or ambiguity, and the framework and coding criteria were refined accordingly to produce the final codebook.

Using the final framework, all three authors independently coded all 293 free-text responses. Inter-coder agreement for primary-theme assignment was assessed on the independently coded data using Fleiss’ kappa (κ). Coding discrepancies were subsequently reviewed and resolved through discussion and consensus. Theme frequencies were summarised descriptively across the complete qualitative dataset. As individual responses could be assigned to more than one theme, themes were not mutually exclusive.

### Ethics approval and consent to participate

The study was designed to gather information on the experiences of caregivers and families affected by CLN2 disease and their understanding of and attitudes toward gene-based therapies. The survey was non-interventional, and participation was voluntary and anonymous. No names, contact details, IP addresses, or other directly identifying information were collected.

Before commencing the survey, participants were provided with written information describing its purpose and the intended use of their responses. Participants were informed that the survey aimed to understand the Batten disease community’s experiences with current treatment options, understanding of gene therapy, and the hopes and concerns of current and bereaved caregivers. The introductory information stated that responses would assist the BDSRA Foundation and potential industry partners in better understanding families’ CLN2 disease journeys and help guide future therapeutic research strategies. The survey was also designed to inform BDSRA Foundation program planning, including education, support, and advocacy initiatives. No separate consent form, question or checkbox was used. After receiving this information, participants elected whether to proceed with the survey; implied informed consent to participate was indicated by voluntary completion and submission of the anonymous survey [see **Supplementary Materials**].

Following completion of the survey, the study protocol was reviewed by Advarra, an independent research ethics review board in the United States (IRB#00000971). The board determined that the protocol (Pro00093968) would have qualified as anonymous survey research not requiring ongoing ethics-board oversight under applicable US regulations (45 CFR 46.104[d][2]).

## RESULTS

### Participant background

A total of 98 survey responses were received from caregivers across 19 countries, with the majority from the United States (58/98; 59.2%) and other predominantly English-speaking countries (Table 1). Most respondents were parents of individuals with CLN2 disease (90.8%, n=89), with the remainder comprising extended family members or caregivers. One respondent was an affected individual with CLN2 disease. Most families reported one affected child (78.6%, n=77) while 21.4% (n=21) reported two affected children. The ages of the individuals with CLN2 disease at the time of the survey ranged from under one year old (n=1) to over 16 years old (n=8). At the time of the survey, 85.7% (n=84/98) of affected individuals were living and 14.3 % (n=14/98) had passed away (Fig. 1A).

**Figure 1.**
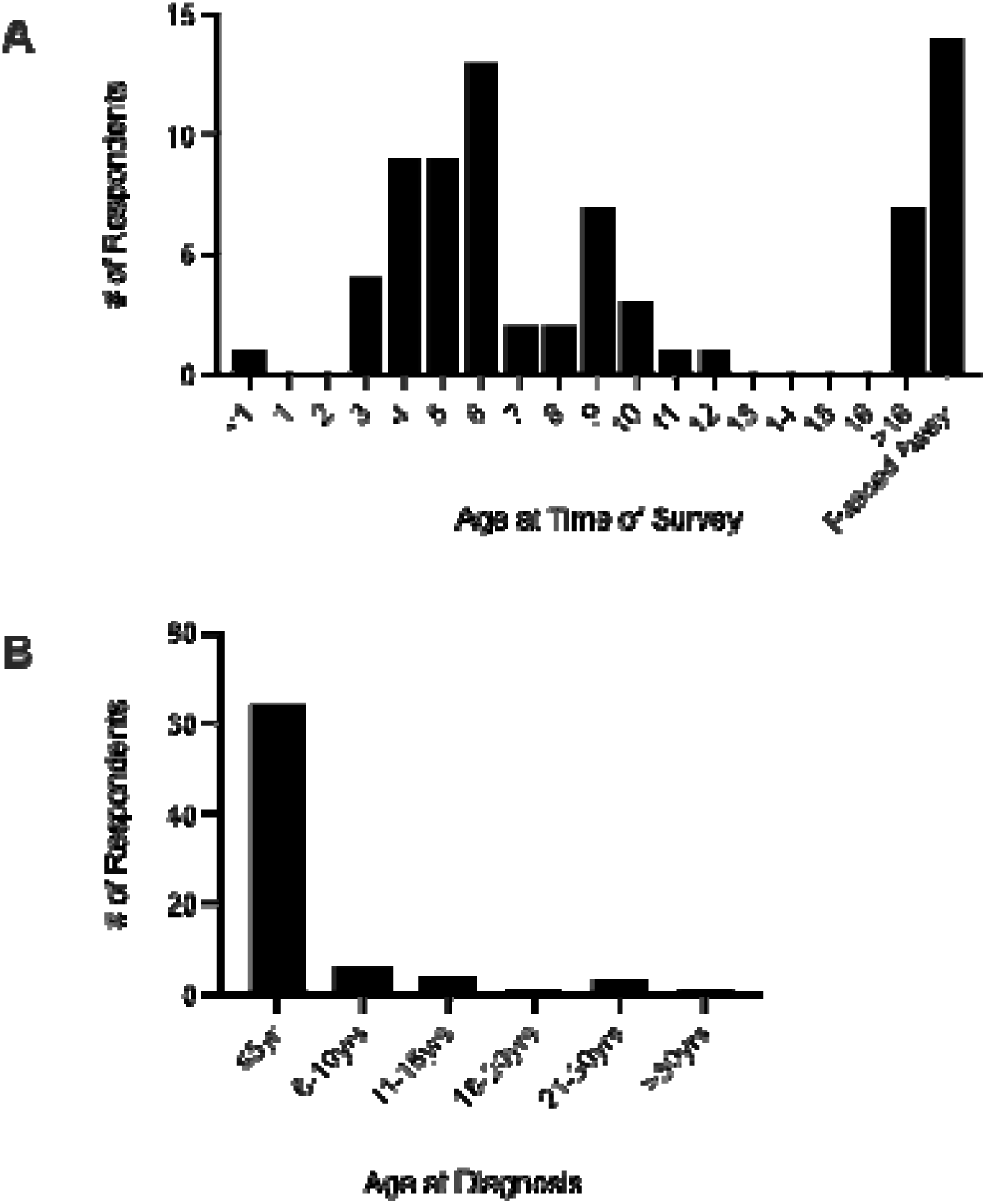
Distribution of age and survival of affected patients at time of survey participation and age at diagnosis as reported by caregiver respondents. A. Of 98 respondents, 85.7% (n=84) indicated the affected individual was living at the time of survey participation, and 14.3% (n=14) indicated the individual had passed. Of the 84 surviving individuals, 69.0% (n=58) were <10 years, and 90.5% (n=76) were younger than 16 years. B. Age at diagnosis was reported by 79 respondents, and ranged from <1 year to 50 years of age, with 64 individuals (81.0%) diagnosed at 5 years or younger, 6 diagnosed at 6-10yrs (7.6%), 4 diagnosed at 11-15yrs (5.1%), one diagnosed at 16-20yrs (1.3%), three diagnosed at 21-30yrs (3.8%). One patient was older than 30yrs (1.3%).

**Table 1.** Geographical distribution of survey responses.

| Country | N<sup>o</sup>. Respondents | # of respondents said Brineura was not available to them or were unsure |
| --- | --- | --- |
| Australia | 6 | 1 |
| Belgium | 3 |  |
| Bosnia Herzegovina | 1 |  |
| Bulgaria | 1 |  |
| Canada | 11 | 1 |
| Croatia | 1 |  |
| Czech Republic | 2 |  |
| Denmark | 3 | 1 |
| Finland | 1 | 1 |
| Georgia | 1 | 1 |
| Germany | 3 |  |
| India | 1 | 1 |
| Ireland | 1 |  |
| New Zealand | 1 | 1 |
| Poland | 1 | 1 |
| Spain | 1 |  |
| United Kingdom | 1 | 1 |
| United States of America | 58 | 4 |
| Zambia | 1 |  |

Age at diagnosis was reported by 79 respondents. Diagnosis most commonly occurred at age 3 years (36.7%, 29/79) or 4 years (31.6%, 25/79). Later diagnoses included six individuals (7.6%) diagnosed between 6–10 years, four (5.1%) between 11–15 years, one (1.3%) between 16–20 years, three (3.8%) between 21–30 years, and one (1.3%) after 30 years of age (Fig. 1B). As survey questions were optional, respondent numbers varied between analyses and denominators are reported for each result.

### Cerliponase alfa treatment experience

Cerliponase alfa was reported as available by 86.7% (85/98) of respondents; 7.1% (7/98) were unsure of its availability and 6.1% (6/98) reported that it was unavailable (Table 1). The survey did not distinguish between regulatory approval, reimbursement/funding, and practical treatment access. Table 1 presents these caregiver-reported responses by country.

Of 79 respondents who answered whether their child had ever received ERT, 72.2% (57/79) reported current or previous treatment and 27.8% (22/79) reported no previous ERT (Fig. 2A). Of the 57 individuals who had received ERT, 94.7% (54/57) were receiving treatment at the time of the survey and 5.3% (3/57) were no longer receiving treatment (Fig. 2B). Treatment was most commonly initiated at age 3 years (42.1%, 24/57) or 4 years (29.8%, 17/57); five individuals (8.8%) commenced treatment before age 3 years and 11 (19.3%) at age 5 years or older (Fig. 2C).

**Figure 2.**
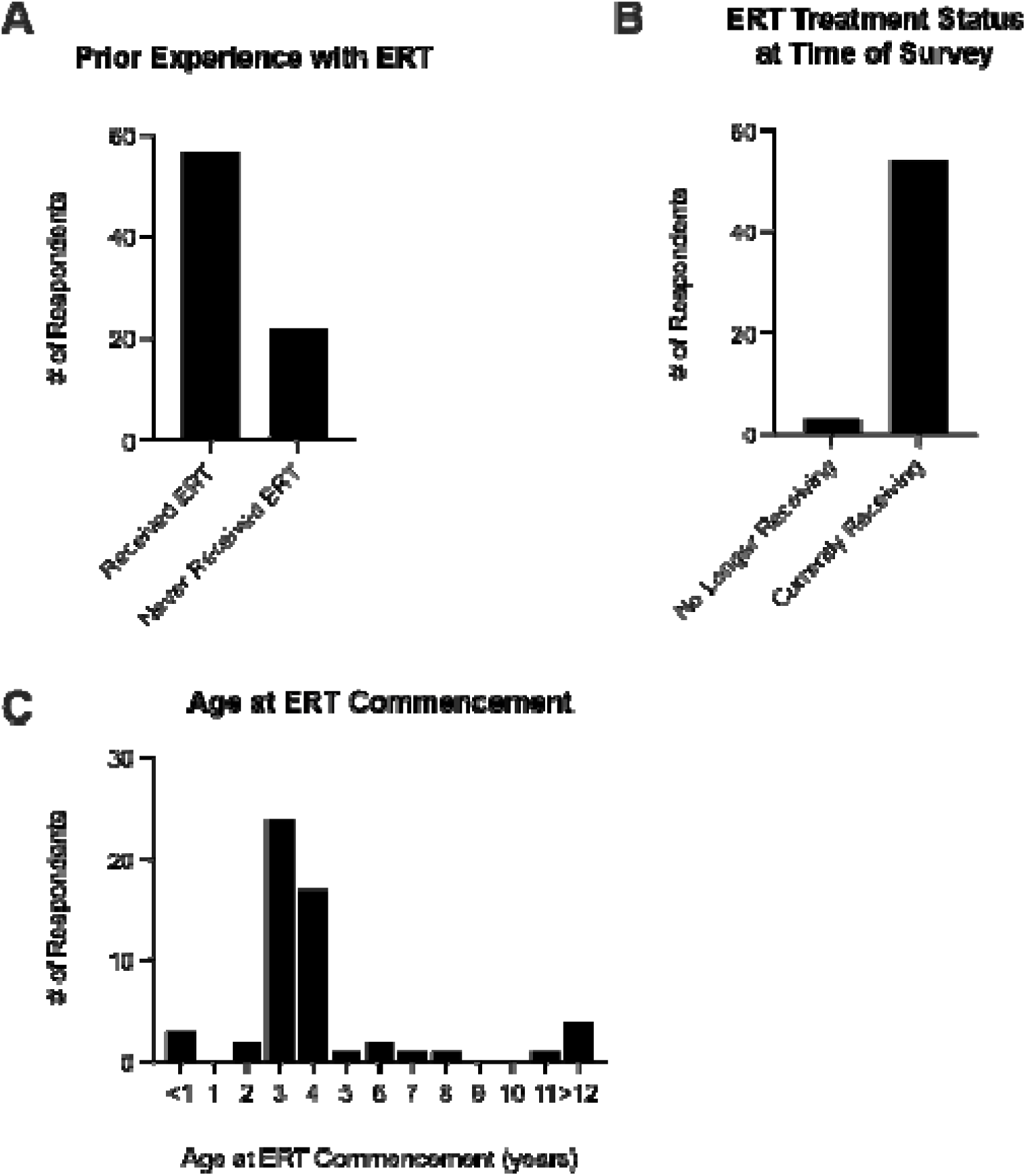
Reported Experience with Enzyme Replacement Therapy. A. 79 participants responded to this question, with 72.2% (n=57) indicating ERT treatment has been received at some time, and 27.9% (n=22) indicated having never received ERT. B. For those 57 participants who had received ERT, 94.7% (n=54) were actively receiving treatment at the time of the survey, 5.3% (n=3) were no longer receiving ERT. C. Age at commencement of treatment, irrespective of whether treatment was current or no longer continuing, was most commonly at 3 years (n=24/57; 42.1%) or 4 years (n= 17/57; 29.8%). Five individuals (8.8%) started before 3 years of age, and 11 individuals (19.3%) began at the age of 5 years or later.

Reported reasons for discontinuation included lack of perceived treatment response (n=1), loss of treatment funding (n=1), and ethical reasons or personal beliefs (n=1). One respondent additionally commented that treatment was “no longer effective.”

Respondents reporting no previous ERT were asked why treatment had not been initiated (Table 2). The most frequently reported reasons were that the affected individual had died before ERT became available (46.1%, 12/26) or was considered too far progressed by the time ERT became available (38.5%, 10/26). The survey did not capture how this determination was made. Other reasons included inability to access treatment funding (7.7%, 2/26), ethical reasons or personal beliefs (3.9%, 1/26), and recent diagnosis with treatment initiation pending (11.5%, 3/26). One participant selected “prefer not to answer.” No respondents indicated that treatment burden outweighed the perceived benefit of ERT.

**Table 2.** Caregiver-reported reasons for not commencing ERT.

| Caregiver-reported reason for not commencing ERT | N<sup>o</sup>. respondents | % |
| --- | --- | --- |
| My child passed before ERT was available | 12 | 46.1 |
| My child was too far progressed when ERT became available | 10 | 38.5 |
| The burden associated with the treatment regimen outweighed the benefit of treatment | 0 | 0 |
| Unable to access funding for treatment (through insurance, government program, etc) | 2 | 7.7 |
| My child has recently been diagnosed, and we are waiting to commence ERT | 3 | 11.5 |
| Ethical reasons/personal beliefs | 1 | 3.9 |
| I prefer not to answer | 1 | 3.9 |
| Other | 4 | 15.4 |

### Caregiver-reported benefits of treatment

Respondents with current or previous ERT experience were asked about benefits they perceived for their affected family member. Fifty-four respondents reported perceived benefits across multiple domains. The most frequently reported benefits were gross motor function, including walking and balance (70.4%, 38/54), and seizure control (70.4%, 38/54), followed by speech and language (42.6%, 23/54), behavioural symptoms (33.3%, 18/54), learning and memory (25.9%, 14/54), and sleep (25.9%, 14/54). Vision benefit was reported less frequently (13.0%, 7/54) (Fig. 3). Several respondents (20.4%, n=11/54) indicated additional benefits, where responses were provided as free text, including “overall comfort/quality of life is improved” (n=1), “emotional, social aspects” (n=1), “tremors” (n=1), “more focused” (n=1) and “receptive language” (n=1).

**Figure 3.**
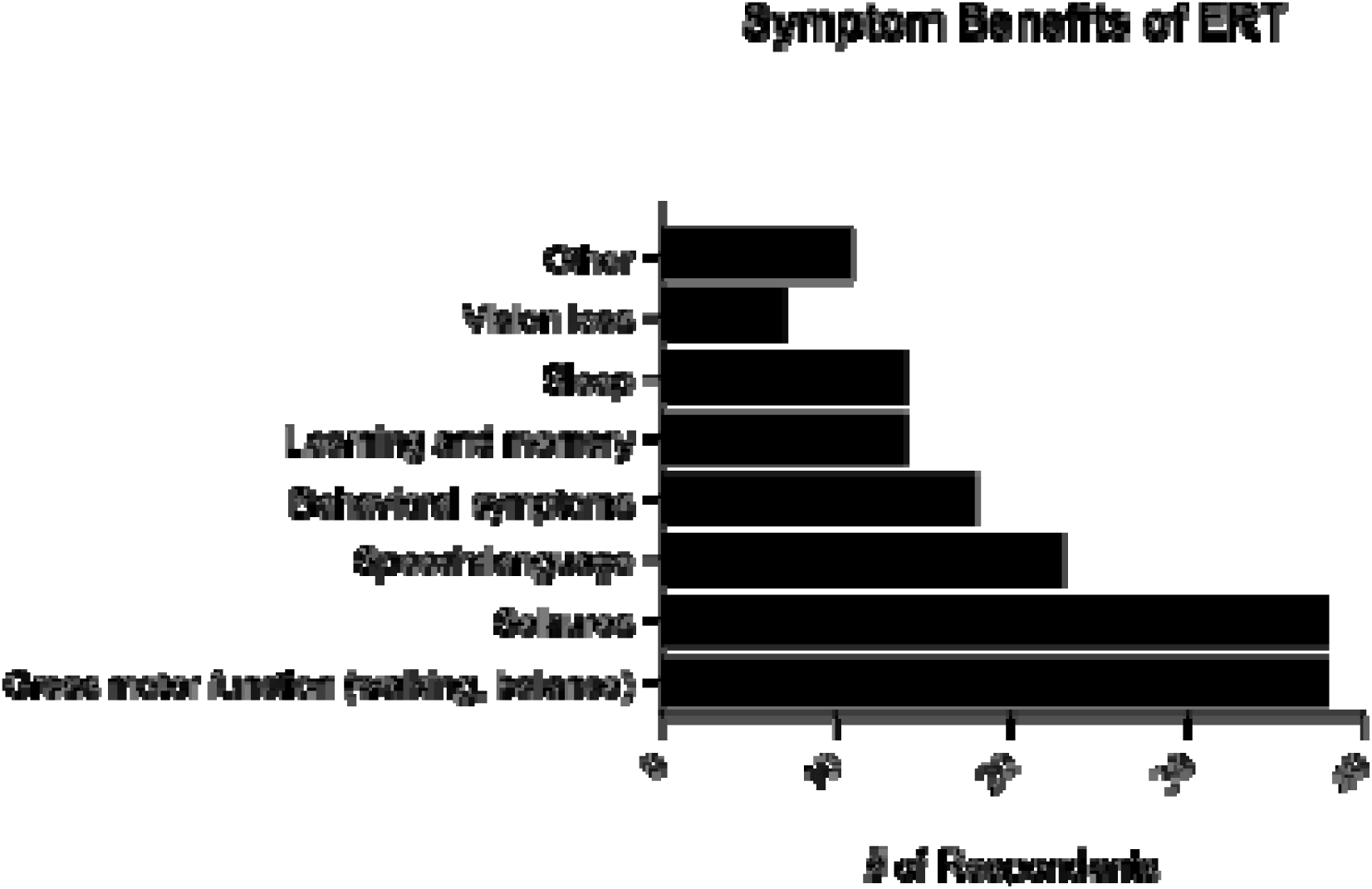
Reported symptom benefit of ERT therapy in CLN2 disease. Based on personal experience, respondents (n = 54) identified CLN2 disease symptoms in their child that they perceived to improve most with ERT. Benefits in gross motor function and seizure control were most frequently reported by caregivers.

When asked whether their current or previous experience with ERT could be improved, 20.4% (11/54) indicated that no improvements were needed. Suggested improvements included changes to infusion dose or frequency (40.7%, 22/54), greater education of hospital staff and healthcare providers (37.0%, 20/54), improvements in infusion delivery methods (33.3%, 18/54), and greater standardisation of procedures relating to ventricular access device placement, access and replacement (31.5%, 17/54). Free-text responses also described the logistical and psychosocial demands of lifelong fortnightly infusions, particularly prolonged hospital visits and travel requirements:

> “*Even though we are grateful, bi-weekly infusions are a lot and exhausting on the child and family*.”
>
> “*Brineura is an amazing and effective tool. I do believe that it can be further optimized*.”
>
> “*I … wish there was a way for us to limit the amount of time that we have to spend in the hospital for these infusions*.”
>
> “*If there was a way to not have a hospital visit all day every 2 weeks.*
>
> *Sedation all day which affects him 3 days after as well.”*

### Treatment burden

Respondents with ERT experience were asked to rate the overall burden of the treatment schedule on a 7-point scale, where 1 represented “no impact at all” and 7 “very impactful.” Sixty-six respondents provided an applicable burden rating, with a mean score of 4.8/7 (median 5; range 1–7) (Fig. 4). Eleven additional respondents selected a score of 0; review of their individual responses confirmed that none had experience with Brineura, and these responses were therefore treated as not applicable and excluded from the calculation.

**Figure 4.**
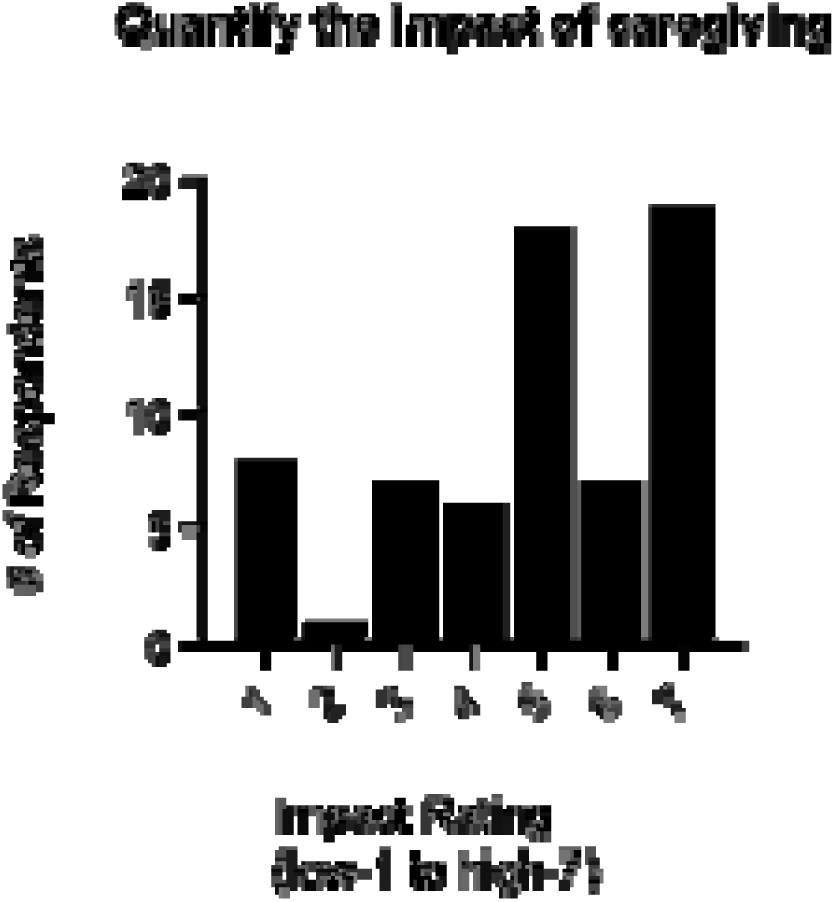
Impact of treatment Respondents were asked to rate the overall burden of the ERT treatment schedule on their family and the affected child on a scale of 1 to 7, where 1 is “No impact at all” and 7 is “Very Impactful.”

### Psychosocial and practical impacts CLN2 disease

When asked which aspects of their lives were affected by caring for an individual with CLN2 disease, emotional and mental stress was the most frequently reported impact (96.1%, 74/77), followed by sleep deprivation (75.3%, 58/77), financial burden (71.4%, 55/77), social isolation (70.1%, 54/77), physical strain (66.2%, 51/77), and travel burden (64.9%, 50/77) (Fig. 5A). Other reported impacts included marriage or relationship challenges (57.1%, 44/77), perceived inability to support family (55.8%, 43/77), difficulty navigating the healthcare system (54.6%, 42/77), taking leave or unpaid leave from work (46.8%, 36/77), reducing work hours (46.8%, 36/77), and leaving employment to become a full-time caregiver (39.0%, 30/77). No respondents indicated that CLN2 disease had no impact on their lives.

**Figure 5:**
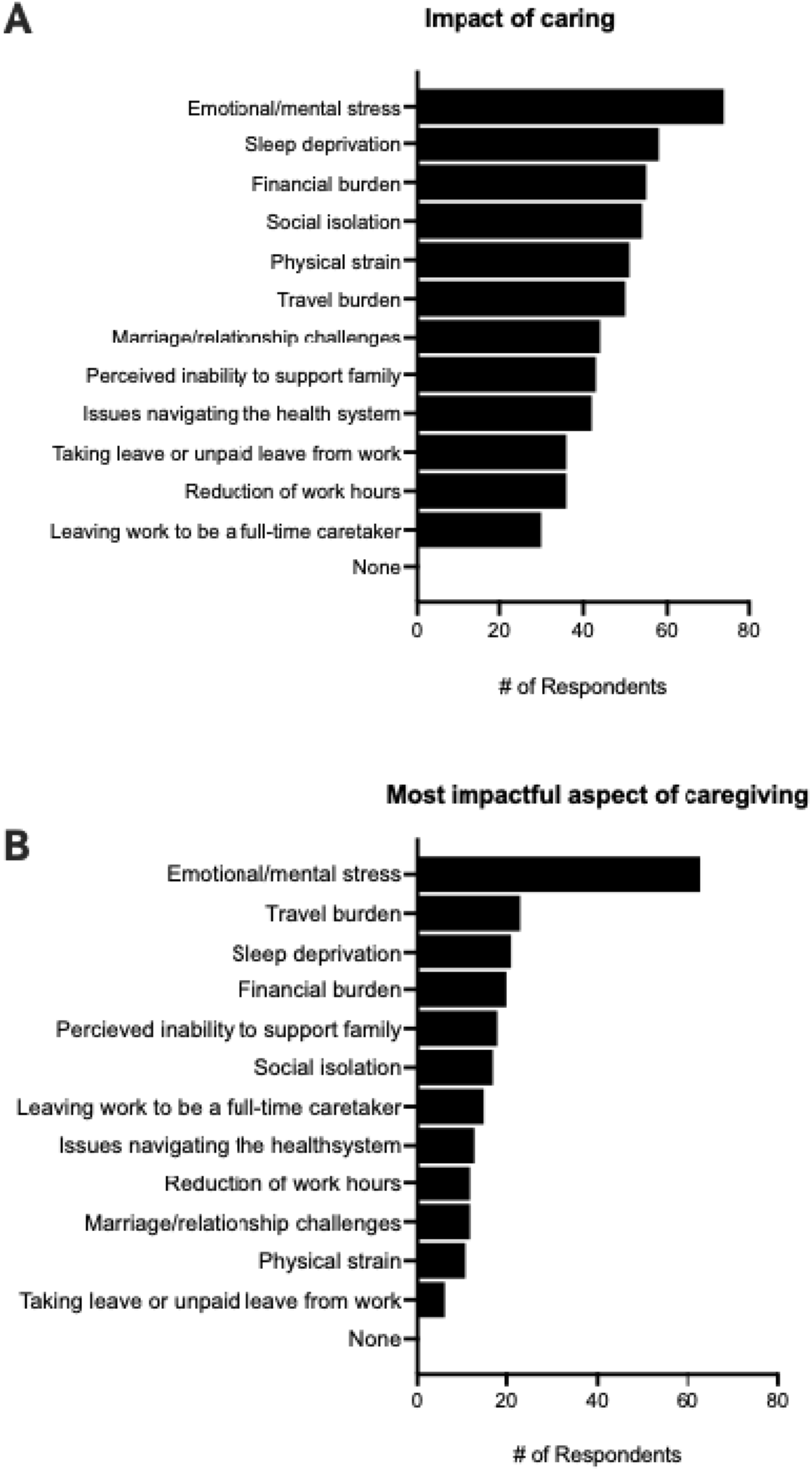
Reported impact of caring for a child with CLN2 disease on personal wellbeing A. Most commonly reported challenges resulting from caring for a child with CLN2 disease. Caregivers (n = 77) were asked to select any impacts of caring for a child with CLN2 disease on employment, family and/or personal well-being. B. Respondents (n = 76) identified up to three factors they considered most impactful/challenging.

When asked to identify up to three of the most challenging impacts, emotional and mental stress was again most frequently selected (82.9%, 63/76), followed by travel associated with treatment and/or other healthcare appointments (30.3%, 23/76), sleep deprivation (27.6%, 21/76), and financial burden (26.3%, 20/76) (Fig. 5B).

### Interest in alternative treatment approaches for CLN2 disease

Most respondents (94.9%, 75/79) indicated a need for additional therapeutic options for CLN2 disease; none indicated that additional therapies were unnecessary, while 5.1% (4/79) were unsure.

Regarding familiarity with gene therapy, 79.2% (61/77) described themselves as extremely, very or somewhat familiar, while 20.8% (16/77) were slightly or not at all familiar. Most respondents (94.8%, 73/77) expressed interest in learning more about gene therapy approaches for CLN2 disease. Preferred sources of information included websites (69.4%, 50/72), Batten disease patient organisations (61.1%, 44/72), the affected individual’s physician (52.8%, 38/72), lay-friendly webinars (48.6%, 35/72), printed materials (48.6%, 35/72), and face-to-face meetings, conferences or educational events (45.8%, 33/72).

When asked about potential participation in an investigational gene therapy trial, 72.4% (55/76) reported that they would be likely or very likely to consider participation. Eight respondents (10.5%) were unsure, one was unlikely, four (5.3%) were definitely unlikely, and eight (10.5%) indicated that their decision would depend on other factors but did not provide specific details (Fig 6).

**Figure 6.**
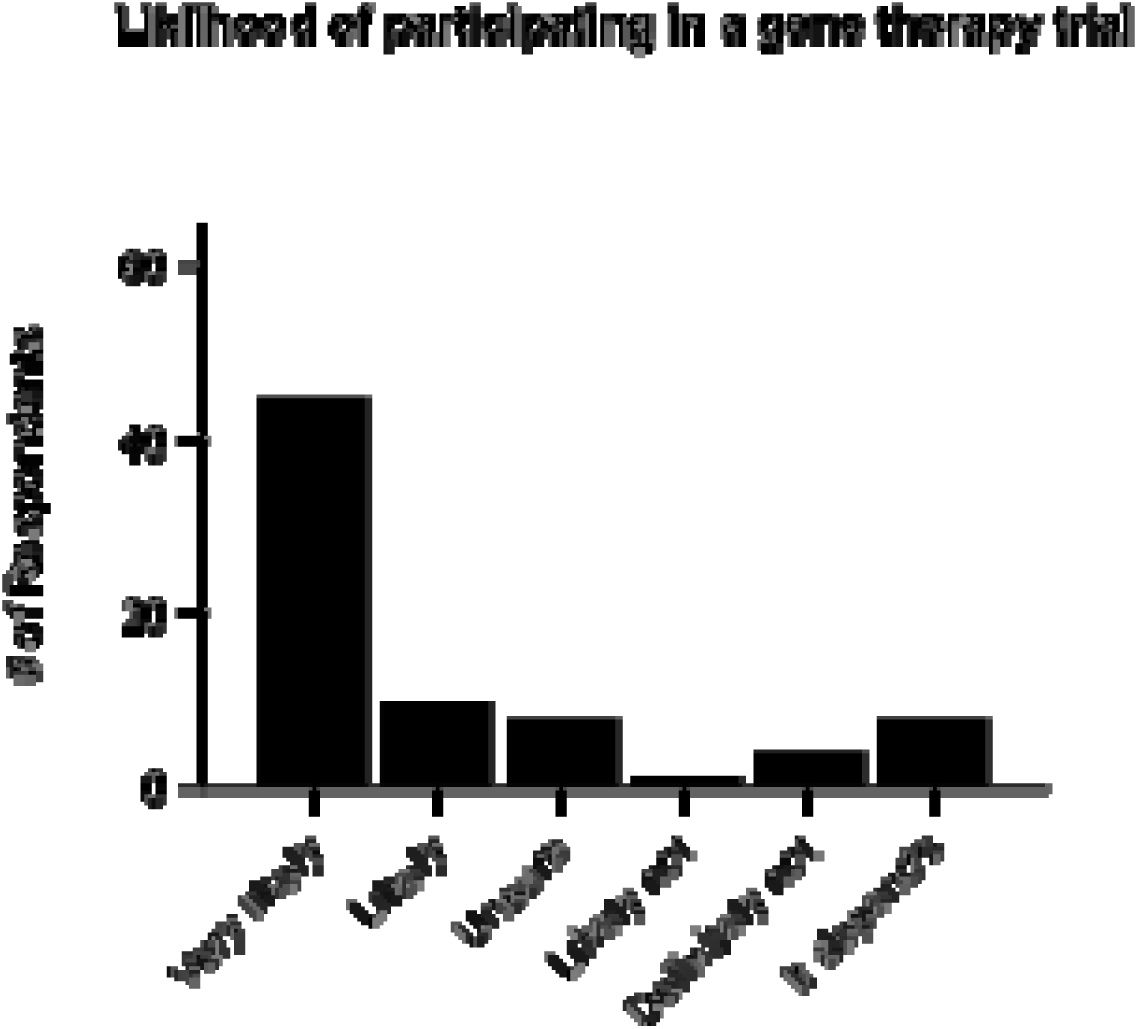
Likelihood to consider participating in a gene therapy clinical trial Respondents were asked how likely it would be for them to consider participating in an investigational gene therapy trial for CLN2 disease, if there was a regulatory-approved research trial available. 72.4% or respondents (n=55/76) indicated they were very likely (45/76) or likely (n=10/76) to consider participating.

### Factors influencing clinical trial participation

Respondents were asked which factors would influence their decision to consider participation in a gene therapy clinical trial, with multiple selections permitted. The most frequently selected considerations were potential safety risks and side effects (57.9%, 44/76), preclinical evidence of safety and efficacy (54.0%, 41/76), and whether ERT would need to be discontinued to participate (54.0%, 41/76). Other considerations included disease progression and the potential for individual benefit (44.7%, 34/76), treatment efficacy (43.4%, 33/76), risks associated with the administration procedure (38.2%, 29/76), potential long-term risks or benefits (35.5%, 27/76), costs associated with participation (34.2%, 26/76), healthcare professional recommendations (27.6%, 21/76), study location and travel logistics (27.6%, 21/76), suitability for participation (26.3%, 20/76), availability of existing treatment options (22.4%, 17/76), burden of ongoing study assessments (17.1%, 13/76), and ethical concerns or personal beliefs (5.3%, 4/76) (Table 3).

**Table 3.** Factors influencing parent/carer decisions regarding participation in a gene therapy clinical trial.

| Factors for consideration | N° Respondents | % |
| --- | --- | --- |
| Potential safety risk and side effects | 44 | 57.9 |
| Preclinical evidence of safety and efficacy (data from cell and animal models) | 41 | 54.0 |
| Will my child need to stop enzyme replacement therapy in order to participate in a study? | 41 | 54.0 |
| Would my child benefit from gene therapy, or are they too far progressed? | 34 | 44.7 |
| Efficacy (will the therapy even work?) | 33 | 43.4 |
| Risks associated with the one-time administration procedure (therapy delivered directly to brain or eyes) | 29 | 38.2 |
| Potential long-term risks or benefits | 27 | 35.5 |
| Costs associated with participating in a study | 26 | 34.2 |
| Recommendations from healthcare providers such as neurosurgeons and neurologists | 21 | 27.6 |
| Study location and travel logistics | 21 | 27.6 |
| Would my child be the best candidate to prove the efficacy of gene therapy, or should treatment be offered to younger/less progressed children first? | 20 | 26.3 |
| Availability of current treatment options | 17 | 22.4 |
| Burden of ongoing assessments for the duration of the study | 13 | 17.1 |
| Ethical concerns/personal beliefs | 4 | 5.3 |
| None | 2 | 2.6 |

### Thematic Analysis of Free-Text Responses

Nine themes were identified across 293 free-text responses from the 10 survey questions included in the thematic analysis (Table 4). Prior to consensus resolution, inter-coder agreement for primary-theme assignment was κ=0.72. Among responses with complete coding by all three authors, at least two coders independently agreed on the primary theme in 95.2% of cases. Coding discrepancies were resolved through discussion and consensus before final theme frequencies were calculated. Individual responses could be assigned to more than one theme, with a maximum of three themes per response; therefore, theme frequencies were not mutually exclusive.

**Table 4.** Thematic analysis of free-text responses.

| Theme | N° | % of |
| --- | --- | --- |

|  | Responses | responses |
| --- | --- | --- |
| Positive experience and benefits of ERT | 16 | 5.5% |
| ERT treatment burden, limitations and need for optimization | 49 | 16.7% |
| Persistent disease burden and unmet therapeutic need | 24 | 8.2% |
| Desire and/or hope for transformative/disease-modifying therapies | 56 | 19.1% |
| Emotional, psychosocial and family impact | 100 | 34.1% |
| Faith, resilience and spirituality as a coping resource | 64 | 21.8% |
| Access and equity in treatment and clinical trials | 5 | 1.7% |
| Regulatory pace and pharmaceutical/ industry transparency | 8 | 2.7% |
| Caregiver and patient voice in research and therapeutic development | 6 | 2.0% |
| Not codable / administrative | 22 | 7.5% |

The most frequently identified theme was emotional, psychosocial and family impact (100/293 responses; 34.1%). Responses described grief, emotional distress, exhaustion and uncertainty associated with the CLN2 disease journey, using words and phrases including *devastating, emotional, exhausting, grief, heartbreaking, life-changing, painful, rollercoaster, stressful, unfathomably heavy, emotionally depleting, life-shattering, not hopeful, sadly optimistic, sceptical*, and *uncertainty* (Fig. 7). One caregiver described the experience as an *“emotional, devastating, journey of grieving for your child while they are still alive.*”

**Figure 7.**
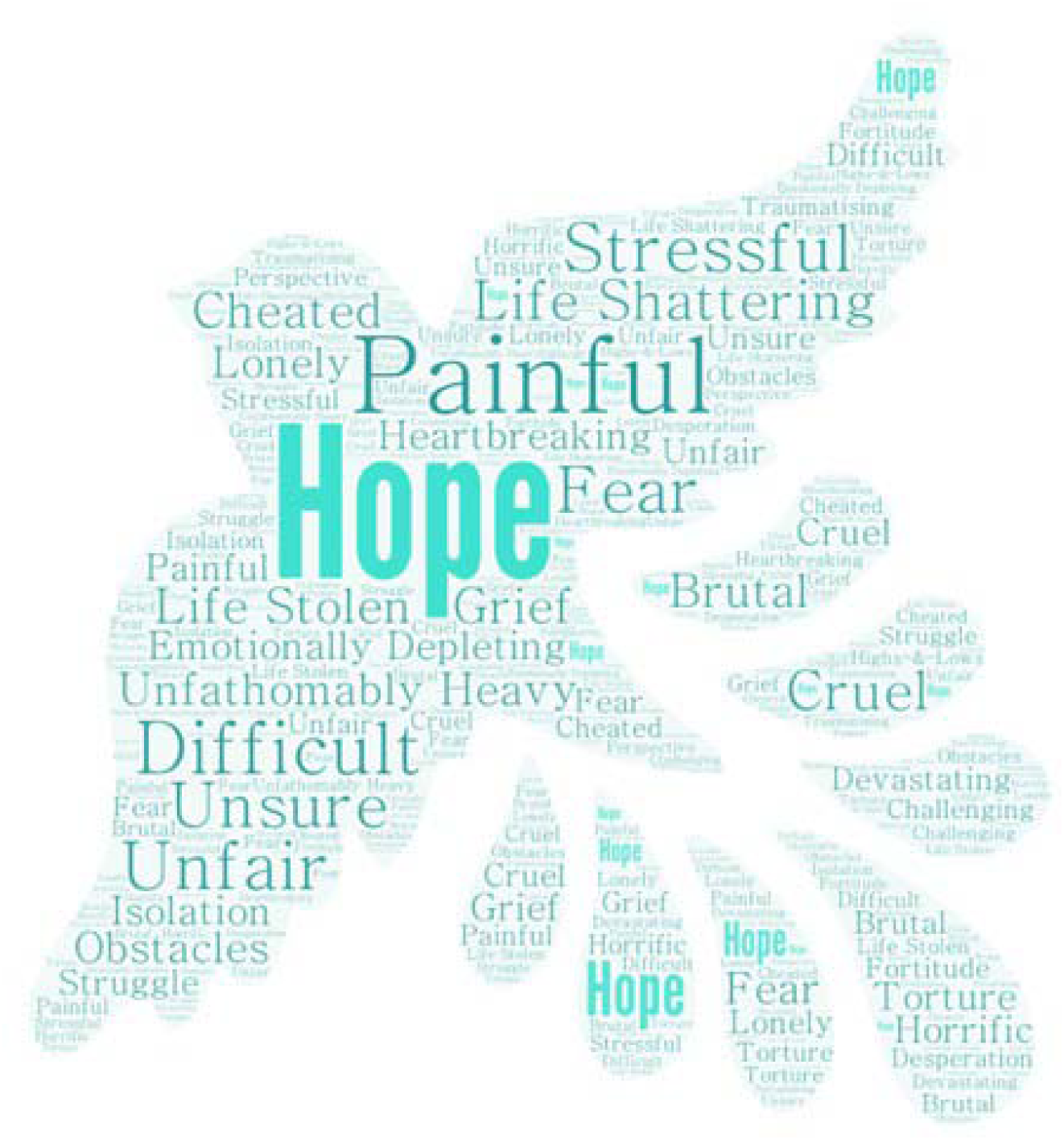
Describing the journey Common words and phrases used by CLN2 caregivers to describe their disease journey.

Faith, resilience and spirituality as a coping resource was also prominent (64/293; 21.8%). Responses within this theme reflected hope, faith and resilience in the context of living with CLN2 disease, using words and phrases including *cautiously optimistic, cure, faith, gene therapy, hope, Jesus, miracle,* and *positive.*

A desire and/or hope for transformative or disease-modifying therapies was identified in 56 responses (19.1%), while 49 responses (16.7%) described ERT treatment burden, limitations and the need for treatment optimisation. A further 24 responses (8.2%) reflected persistent disease burden and unmet therapeutic need. Positive experiences and perceived benefits of ERT were identified in 16 responses (5.5%).

Less frequently identified themes included regulatory pace and pharmaceutical/industry transparency (8/293; 2.7%), caregiver and patient voice in research and therapeutic development (6/293; 2.0%), and access and equity in treatment and clinical trials (5/293; 1.7%). Responses within these themes described concerns regarding the pace of therapeutic development and regulatory processes, transparency from pharmaceutical companies, differences in treatment access between countries, and access to clinical trials for individuals with non-classical or “atypical” CLN2 disease. One caregiver commented, “*We just wish the pharmaceutical companies were more transparent with us […] so our family can have a better understanding as to what to expect.”*

## DISCUSSION

This study provides important insights into the lived experiences of families affected by CLN2 disease in the era of disease-modifying treatment. To our knowledge, it represents the largest caregiver survey conducted specifically in CLN2 disease and the first to examine caregiver perspectives on both current ERT and emerging gene-targeted therapies. Three principal findings emerged: caregivers perceived cerliponase alfa as providing meaningful clinical benefit; substantial disease, treatment and caregiver burden persisted despite treatment; and there was strong interest in additional therapeutic approaches, particularly gene-targeted therapies.

### Treatment benefits and limitations

Caregivers most frequently reported benefits in gross motor function, seizure control and speech and language. The reported motor and language benefits are consistent with clinical evidence demonstrating sustained slowing of decline in these domains [16,19]. Improved seizure outcomes have also been reported with treatment [22,26,27], together with improved health-related quality of life [28], extended survival [19,22] and overall clinical stability [24]. Evidence that treatment initiated presymptomatically can delay the onset of clinical manifestations [23], further supports the importance of early diagnosis and timely access to treatment, including consideration of CLN2 disease for newborn screening programs [22,24,25,50].

A small proportion of caregivers reported perceived benefit in vision despite evidence that retinal degeneration continues during ICV cerliponase alfa treatment [31,32]. This may reflect younger age or earlier disease stage before substantial retinal decline which typically occurs from 48-72 months [31,32], variability in disease trajectory, or differences between caregiver-perceived visual function and objectively measured retinal degeneration. These findings should therefore not be interpreted as evidence of an effect of ICV ERT on retinal disease.

### Treatment burden remains substantial

Despite perceived clinical benefits, ERT was associated with considerable treatment burden, with a mean burden score of 4.8/7. Free-text responses similarly described the logistical and psychosocial demands of lifelong fortnightly ICV infusions, including prolonged hospital visits, travel requirements, and disruption to family life. Notably, however, no respondents who had not commenced ERT identified treatment burden as a reason for non-initiation.

These findings illustrate an important distinction between treatment benefit and treatment burden: caregivers may perceive substantial benefit from ERT while simultaneously experiencing considerable burden associated with its administration. This has implications for future therapeutic approaches that aim not only to improve disease control but also to reduce the frequency, invasiveness, and practical demands of treatment.

### Caregiver and family burden

Beyond treatment administration, caregivers reported substantial effects of CLN2 disease across multiple aspects of family life. Emotional and mental stress was the most frequently reported impact, followed by sleep deprivation, financial burden, social isolation, physical strain, and travel associated with treatment and other healthcare needs. These findings are consistent with previous literature demonstrating substantial caregiver burden in Batten disease and other lysosomal storage disorders [42–45,49].

Together, these data suggest that while ERT has transformed the clinical trajectory of CLN2 disease, treatment success must be considered not only in terms of neurological outcomes but also caregiver impact and treatment feasibility.

### Caregiver burden

Beyond treatment logistics, caregivers reported substantial broader impacts associated with caring for a child with CLN2 disease. The most frequently reported challenges were emotional and mental stress, sleep deprivation, travel associated with healthcare needs, and financial strain. These findings align with previous literature demonstrating significant caregiver burden in Batten disease and other lysosomal storage disorders [42–45,49].

The thematic analysis provides further context to these quantitative findings. Emotional, psychosocial and family impact was the most frequently identified theme across the 293 free-text responses. Expressions of grief, exhaustion, distress and uncertainty suggest substantial psychological burden throughout the disease course, including anticipatory grief associated with progressive loss of function and reduced life expectancy. At the same time, faith, resilience and spirituality emerged as a prominent theme, with expressions of hope and cautious optimism frequently accompanying descriptions of considerable emotional burden.

Together, these findings demonstrate that the impact of CLN2 disease extends beyond the affected individual to the wider family. This is consistent with published CLN2 clinical management guidelines emphasising multidisciplinary care that addresses both clinical manifestations and the psychosocial needs of families and caregivers [41,47]. The findings further support caregiver psychosocial assessment and support as components of comprehensive CLN2 disease management and consideration of caregiver and family burden in clinical care and future research.

### Implications for future therapies

The high proportion of respondents indicating a need for additional therapeutic options (94.9%) and interest in learning more about gene therapy (94.8%) demonstrates substantial interest in emerging treatments. Almost three-quarters of respondents (72.4%) reported that they would be likely or very likely to consider participation in an investigational gene therapy trial. These findings align closely with the research priorities identified by Augustine et al. [50], in which development of effective gene-targeted therapies emerged as the highest-ranking research priority for the Batten disease community.

Importantly, interest in gene therapy was accompanied by careful consideration of potential risks and benefits. The most frequently selected factors influencing potential trial participation were safety risks and side effects, preclinical evidence of safety and efficacy, and whether existing ERT would need to be discontinued. Disease stage and the potential for individual benefit, treatment efficacy, procedural risks, long-term risks and benefits, and the financial and logistical demands of trial participation were also important considerations. These findings suggest that families are highly interested in gene-targeted approaches while carefully weighing their potential benefits against safety, existing treatment options, disease stage, and study-related burden.

The thematic analysis further identified a strong desire and hope for transformative or disease-modifying therapies. This finding is particularly relevant given the limitations of current ERT identified by respondents, including the burden of lifelong fortnightly administration and the ongoing need for therapies capable of addressing manifestations not adequately treated by ICV ERT, particularly progressive retinal degeneration. Together with the research priorities identified by Augustine et al. [50], these findings demonstrate substantial community interest in therapeutic approaches capable of providing broader and more durable disease modification.

### Future therapeutic directions

Gene-targeted therapies represent an important area of therapeutic development for CLN2 disease. Adeno-associated virus (AAV) vectors have emerged as promising platforms for delivery of functional genes to the CNS. Preclinical studies demonstrate that AAV-mediated gene transfer can restore TPP1 expression in CLN2 disease models. Because TPP1 can be distributed between cells through cross-correction, complete CNS transduction may not be necessary to achieve therapeutic benefit. Achieving sufficient enzyme expression may therefore have the potential to meaningfully modify disease progression following a single administration [48]. However, AAV-based approaches also present challenges, including dose-related safety considerations, potential off-target effects, and complex manufacturing requirements that may influence cost and scalability.

Several investigational gene therapy programs for CLN2 disease are currently in development. Early clinical studies of intraparenchymal AAV delivery have demonstrated increased TPP1 expression and signals of slowed neurodegeneration, although disease progression was not fully halted [51]. Additional CNS- and ocular-directed approaches are advancing in parallel, with early clinical data suggesting acceptable safety profiles and potential neurological and retinal benefits [38,52]. Collectively, these efforts reflect growing momentum toward therapies aimed at addressing the underlying genetic cause of CLN2 disease.

Regulatory agencies increasingly recognise the importance of incorporating patient and caregiver perspectives into therapeutic development to ensure that clinical trial endpoints and outcome measures reflect aspects of disease and treatment that are meaningful to affected communities. The findings from this survey provide insight into caregiver priorities, including reducing treatment burden, preserving function, improving quality of life, and carefully considering the safety and study-related burdens associated with investigational therapies. These perspectives may inform future clinical trial design by helping to identify acceptable treatment and study-related burdens, prioritise functional and quality-of-life outcomes that are meaningful to patients and families, and guide the selection of patient-and caregiver-relevant endpoints and outcome measures. Continued development of gene-targeted therapies has the potential not only to further modify disease progression, but also to address important unmet needs identified by families, including reducing treatment burden and improving long-term outcomes for individuals living with CLN2 disease and their families.

### Study limitations

This study has several limitations. Participants were recruited through patient advocacy networks, which may introduce selection bias toward families who are more highly engaged with patient organisations. The survey was conducted in English only, and the sample was weighted toward the United States (58/98 respondents; 59.2%) and other predominantly English-speaking, high-resource settings. Although responses were received from 19 countries, the cohort should therefore not be considered globally representative, and perspectives from non-English-speaking and lower-resource settings may be underrepresented. Differences in healthcare systems, treatment and clinical trial access, and attitudes toward emerging therapies may limit the generalisability of some findings. This may be particularly relevant to non-classical or “atypical” CLN2 phenotypes, which have been reported at higher frequencies in some populations underrepresented in this survey.

Nevertheless, the geographical breadth and size of the cohort provide valuable insight into the experiences of a substantial and diverse group of families affected by this ultra-rare disease.

The survey relied on self-reported data that were not clinically verified. The survey also did not distinguish between regulatory approval, reimbursement or funding, and practical access to treatment. These are distinct determinants of treatment availability, and approval of cerliponase alfa within a jurisdiction does not necessarily mean that treatment is funded, accessible at an appropriate specialist centre, or available to an individual patient. Caregiver-reported treatment availability should therefore be interpreted as reflecting families’ experience of access rather than formal regulatory or reimbursement status.

Finally, the study was not designed or powered to compare clinically relevant subgroups, including treated versus untreated individuals or classical versus non-classical CLN2 disease. Future studies should prospectively recruit sufficient numbers across these populations to enable meaningful comparisons. Although one affected individual participated, the study primarily reflects caregiver perspectives; greater direct participation from individuals living with CLN2 disease, particularly those with later-onset and non-classical phenotypes, would provide valuable additional insight.

## CONCLUSION

Despite meaningful advances with cerliponase alfa, CLN2 disease continues to impose substantial disease, treatment, and caregiver burden on affected individuals and their families. While ERT has significantly altered the natural history of the disease, its limitations – including ongoing retinal degeneration and the demands of lifelong invasive treatment – demonstrate the need for therapies that can provide broader disease modification while reducing treatment burden.

This caregiver survey demonstrates substantial interest in emerging gene-targeted therapies, while also identifying safety, evidence of efficacy, preservation of function, whether established ERT would need to be discontinued, and treatment- and study-related burden as important considerations for families. These findings support continued development of therapeutic approaches capable of more comprehensively addressing both CNS and ocular manifestations of CLN2 disease while improving the treatment experience for affected individuals and their families.

As the therapeutic landscape continues to evolve, incorporating patient and caregiver perspectives into therapeutic development and clinical trial design will be important to ensure that new approaches address outcomes that are meaningful to those living with CLN2 disease.

Future progress in CLN2 disease will depend not only on scientific advances, but on ensuring that new therapies meaningfully reduce disease burden and address the outcomes that matter most to affected families.

## Data Availability

All data produced in the present study are available upon reasonable request to the authors.

## DECLARATIONS

### Ethics Approval and Consent to Participate

Following completion of the study, study protocol was reviewed by Advarra (IRB#00000971) per the US regulations at 21 CFR 50, 56. The board determined protocol (Pro00093968) would have met criteria for Exemption from IRB oversight under 45 CFR 46.104(d)(2).

The study was non-interventional in nature, and participants completed the survey voluntarily and anonymously. No names, contact details, IP addresses, or other identifying information were collected. All participants were provided with study information prior to survey commencement. Implied informed consent to participate was obtained through voluntary completion and submission of the anonymous survey.

### Clinical trial number

Not applicable.

### Consent for publication

Not applicable.

### Availability of data and materials

The datasets used and/or analyzed during the current study are available from the corresponding author upon reasonable request.

### Competing Interests (from each author)

IW is Head of Research and Medical Affairs for the BDSRA Foundation and BDSRA Australia and was involved in survey design, data analysis and manuscript preparation. JC, KV and GY are employees of Latus Bio and were involved in survey design, data analysis and manuscript preparation. Latus Bio provided funding for a medical writer to assist with manuscript preparation.

AFP served as President and CEO of the BDSRA Foundation and was involved in survey design.

Clinician authors [RAN and AS] have received consultancy fees from Latus Bio.

Each author has completed the ICMJE declaration forms, none of which indicated any competing interests that may have influenced the validity of this research. Copies of the completed ICMJE forms are available upon reasonable request.

### Funding Partners

Latus Bio provided funding support for the IRB Exemption review and for a medical writer to assist with this manuscript. Medical writing and editorial support were provided by Heather Mason, PhD, of Coufetery Communications, including assistance with manuscript preparation, editing, formatting, and incorporation of author comments, and were funded by Latus Bio.

### Authors’ Contributions

Conceptualization: I W, GY. Methodology: IW, GY, AFP, CS, JC. Data curation: I W, GY, KV, CS. Data analysis: IW, GY, KV, JC, RAN, AS. Figures preparation: GY, KV, IW. Project administration: I W, GY. Visualization: I W, GY. Writing – original draft: I W, GY, KV. Writing – review & editing: All authors. Each author has approved the submitted version, along with supporting documentation, and has agreed both to be personally accountable for the authors’ own contributions and to ensure that questions related to the accuracy or integrity of any part of the work, even ones in which the author was not personally involved, are appropriately investigated, resolved, and the resolution documented in the literature.

## Acknowledgments

The authors wish to thank the CLN2 community for providing their candid insight into their lived experiences, challenges, and hopes for future treatment options for CLN2 disease. Our gratitude to Heather L. Mason at Coufetery Comms, France, for medical writing support, and to Erika Augustine MD for constructive comments on the manuscript.

