## Supplementary Materials for "Understanding the CLN2 Disease Journey: A Survey of Caregiver Experiences, Treatment Burden, and Treatment Perspectives"

CLN2 Batten Disease Caregiver Survey: Understanding the Patient Journey, Treatment Approaches and Potential Therapeutic Opportunities for CLN2 Diseas

**This survey is intended for all current and bereaved parents of children and young people aﬀected by CLN2 disease. Your insights and experiences as a caregiver for an aﬀected child, or multiple aﬀected children, are invaluable in helping us to understand and ultimately help improve the standards of care and treatment options for patients. The goal of this survey is to gain insight into the Batten disease community’s experience with current treatment options, the overall understanding of gene therapy, and the hopes and concerns of current and bereaved caregivers when thinking about gene therapy as a possible treatment for CLN2 disease. The responses you provide through this survey will assist the BDSRA Foundation and potential industry partners in better understanding your family’s CLN2 disease journey and help guide future therapeutic research strategies.**

**Signiﬁcant advances have been made in the treatment of CLN2 disease with intraventricular Cerliponase alfa enzyme replacement therapy (hereafter referred to as ERT) now widely available throughout the world. However, additional therapy options continue to be explored that may potentially decrease current treatment burden, halt disease progression, or even prevent disease onset when treated pre-symptomatically. One approach being considered by several biotechnology companies involves the use of viral vector-mediated gene therapy.**

**Gene therapy is a cutting-edge medical approach that aims to treat medical conditions by introducing genetic material, such as a small segment of DNA or a gene, into the cell nucleus. Gene therapies often involve the use of viruses (modiﬁed so they do not cause disease) as delivery vehicles for ‘healthy’ copies of a missing or defective gene. In the case of CLN2 disease, administration of these gene-carrying viruses directly into the brain and retina of patients may be required. It is hypothesized that gene therapy could be a one-time treatment to express the missing TPP1 gene over the long-term and potentially alter the course of CLN2 disease, reducing or eliminating the need for reoccurring, life-long treatments.**

**No personally identiﬁable information will be collected in this survey. We request that for multiple aﬀected children, you ﬁll in one survey per child.**

**We sincerely thank you in advance for taking the time to participate in this important survey. If you have any questions, please feel free to reach out to us anytime.**

**The ﬁnal day to complete the survey will be March 29, 2024. Yours in hope,**

**Amy Fenton-Parker**

**CEO and President, BDSRA Foundation**

**Ineka Whiteman, PhD**

**Head of Research and Medical Aﬀairs, BDSRA Foundation**

#### About CLN2 disease:

***CLN2 disease is a genetically inherited lysosomal storage disorder, in which aﬀected individuals lack an enzyme called tripeptidyl peptidase 1 (TPP1) that breaks down molecules such as fats and proteins in compartments within the body’s cells called lysosomes. Currently, the only approved therapy for CLN2 disease is intraventricular Cerliponase alfa enzyme replacement therapy (ERT) that delivers the missing TPP1 enzyme directly to the brain of children with CLN2 disease. This therapy requires that patients receive ERT brain infusions every two weeks in a hospital setting.***

Demographic Information

CLN2 Batten Disease Caregiver Survey: Understanding the Patient Journey, Treatment Approaches and Potential Therapeutic Opportunities for CLN2 Diseas

### 1. What is your country of residence?

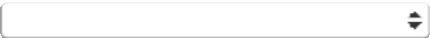

### * 2. Number of aﬀected children (living or deceased)

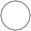
 1

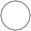
 2

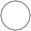
 3

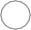
 >3

### 3. Your relationship to the aﬀected child:

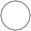
 Parent
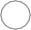
 Guardian

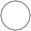
 Other (please specify)

### 4. Aﬀected child's age at time of survey:

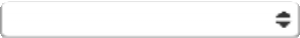

### 5. Is enzyme replacement therapy (Cerliponase alfa) currently available for the treatment of CLN2 disease in your country of residence?

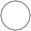

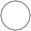
 Yes
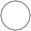
 No

Unsure

Experience with the currently available treatments for CLN2 disease

CLN2 Batten Disease Caregiver Survey: Understanding the Patient Journey, Treatment Approaches and Potential Therapeutic Opportunities for CLN2 Diseas

### 6. How old was your child when they were diagnosed with CLN2 disease?

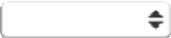

### 7. Has or did your aﬀected child ever receive enzyme replacement therapy?

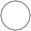
 Yes

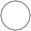
 No (skip ahead to Question 13)

### 8. How old were they (in years) when enzyme replacement therapy ﬁrst commenced?

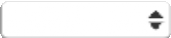

### 9. Is your child currently receiving enzyme replacement therapy?

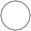
 Yes (skip to question 11)
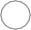
 No

### 10. If you answered "no" to Question 9, please indicate why your child is no longer receiving enzyme replacement therapy. Please select ALL that apply.

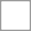
 No longer able to access funding for treatment (through insurance, government program, etc)
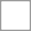
 Ethical reasons/personal beliefs

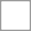
 My child did not respond to treatment

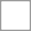
 The side eﬀects of treatment outweighed the beneﬁt of the treatment for my child/ren
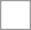
 My child passed away

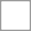
 Other (please provide further comment in text box below)
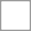
 I prefer not to answer

Other (please specify)

### 11. If your child is receiving/has received enzyme replacement therapy, which of the following symptoms of CLN2 disease do/did you feel the therapy provides most beneﬁt for in your child, based on your own personal experience? Please select ALL that apply.

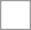
 Gross motor function (walking, balance)
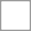
 Speech/language

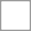
 Vision loss

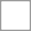
 Behavioral symptoms
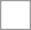
 Learning and memory
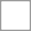
 Seizures

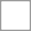
 Sleep

Other (please specify)

### 12. Do you feel your current/previous experience with enzyme replacement therapy could be improved? If so, in what way? (Select ALL that apply)

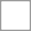
 More hospital/healthcare provider education
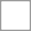
 Improvements in delivery methods

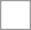
 More standardized procedures, such as with port placement/access/replacement, etc.
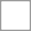
 Change in infusion/dosing frequency (please provide further comment below)

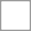
 I do not feel it could be improved.

Other (please specify)

### 13. If your child has never received enzyme replacement therapy, please indicate why (select ALL that apply).

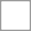
 My child passed before enzyme replacement therapy was available

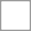
 My child was too far progressed when enzyme replacement therapy became available
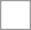
 The burden associated with the treatment regimen outweighed the beneﬁt of treatment
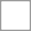
 Unable to access funding for treatment (through insurance, government program, etc)
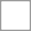
 My child has recently been diagnosed and we are waiting to commence ERT

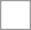
 Ethical reasons/personal beliefs
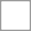
 I prefer not to answer

Other (please specify)

### * 14. Now that enzyme replacement therapy is available to treat individuals with CLN2 disease, do you feel there is still a need for alternative treatments to be developed for CLN2 disease? Why or why not?

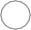
 Yes
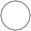
 No

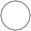
 Unsure

Please provide any comments below.

Carer Burden and Impact

CLN2 Batten Disease Caregiver Survey: Understanding the Patient Journey, Treatment Approaches and Potential Therapeutic Opportunities for CLN2 Diseas

* 15. If your child has ever received enzyme replacement therapy, on a scale of 1 to 7, where 1 is "No impact at all" and 7 is "Very Impactful," **based on your own personal experience, how would you rate the overall burden the treatment schedule places on you, your family and the aﬀected child?**

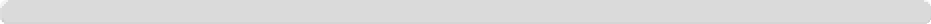

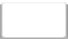

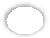

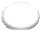

1, No impact at all

7, Very impactful

* 16. In terms of the impact of **caring for a child with CLN2 disease** has had on your employment, family and/or personal well-being. Please select ALL that apply.

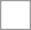
 Financial burden

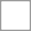
 Had to take unpaid leave/vacation time to care for child

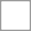
 Had to reduce hours at job/take pay cut for job with less responsibilities
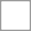
 Had to leave paid work to care full time

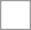
 Travel associated with treatment and/or other healthcare appointments
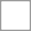
 Emotional/mental stress

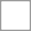
 Physical strain
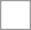
 Social isolation

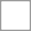
 Issues navigating the healthcare system
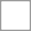
 Sleep deprivation

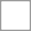
 Feeling you are not meeting other family members' needs
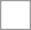
 Marriage/relationship challenges

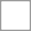
 None

Other (please specify)

### 17. Which of these impacts has been the most challenging? (SELECT UP TO 3)

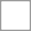
 Financial burden

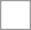
 Having to take unpaid leave/vacation time to care for child

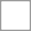
 Having to reduce hours at job/take pay cut for job with less responsibilities
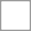
 Had to leave paid work to care full time

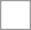
 Travel associated with treatment and/or other healthcare appointments
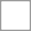
 Emotional/mental stress

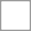
 Physical strain
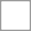
 Social isolation

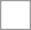
 Issues navigating the healthcare system
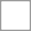
 Sleep deprivation

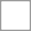
 Feeling I am not meeting other family members' needs
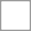
 Marriage/relationship challenges

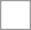
 None

Other (please specify)

General Understanding and Attitudes Toward Gene Therapy

CLN2 Batten Disease Caregiver Survey: Understanding the Patient Journey, Treatment Approaches and Potential Therapeutic Opportunities for CLN2 Diseas

### * 18. What phrase best describes your general understanding and awareness of gene therapy for treating disease?

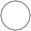
 Extremely familiar
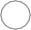
 Very familiar

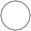
 Somewhat familiar
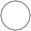
 Slightly familiar
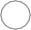
 Not at all familiar

### 19. Would you be interested in learning more about gene therapy approaches to treat CLN2 disease?

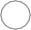

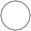
 Yes No

### 20. How would you like to receive information about gene therapy? Please select ALL that apply.

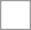
 Websites
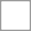
 Television

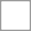
 Lay-friendly webinar (e.g., Zoom, Microsoft Teams)
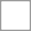
 Print materials (books, papers, magazines, pamphlets)
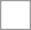
 Face-to-face meetings/conferences/educational events
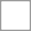
 From my child's physician

From my local Batten's disease patient organization

Considerations for CLN2 Gene Therapy Clinical Trial Participation

CLN2 Batten Disease Caregiver Survey: Understanding the Patient Journey, Treatment Approaches and Potential Therapeutic Opportunities for CLN2 Diseas

### * 21. If there was an investigational gene therapy treatment approach to treating CLN2 disease in a regulatory-approved clinical research setting, how likely would you be to consider participating?

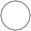
 Very likely
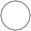
 Likely

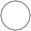
 Unsure
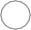
 Likely not

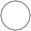

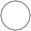
 Deﬁnitely not It depends

### * 22. What factors would inﬂuence your decision to enroll your child in a gene therapy clinical study? Please select ALL that apply.

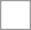
 Potential safety risk and side eﬀects

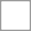
 Preclinical evidence of safety and eﬀicacy (data from cell and animal models)

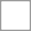
 Risks associated with the one-time administration procedure (therapy delivered directly to brain or eyes) Will my child need to stop enzyme replacement therapy in order to participate in a study?

Would my child beneﬁt from gene therapy or are they too far progressed?

Would my child be the best candidate to prove the eﬀicacy of gene therapy or should treatment be oﬀered to younger/less progressed children ﬁrst?

Recommendations from healthcare providers such as neurosurgeons and neurologists Costs associated with participating in a study

Burden of ongoing assessments for the duration of the study Ethical concerns/personal beliefs

Eﬀicacy (will the therapy even work?) Potential long-term risks or beneﬁts

Availability of current treatment options Study location and travel logistics

None

Other (please specify)

Additional comments

CLN2 Batten Disease Caregiver Survey: Understanding the Patient Journey, Treatment Approaches and Potential Therapeutic Opportunities for CLN2 Diseas

### 23. What word or phrase describes your family's journey with Batten disease?

### 24. What word or phrase best describes your family’s hope for the future?

25. **Optional:** Do you have any additional comments, concerns, or suggestions related to gene therapy clinical trials or the current standard of care for CLN2 Batten disease that you would like to share?
